# Monitoring of anti-SARS-CoV-2 IgG antibody immune response in infected and immunised healthcare workers in Hungary: a real-world longitudinal cohort study

**DOI:** 10.1101/2021.05.16.21257288

**Authors:** Judit Gervain, Katalin B. Szabó, Erika H. Baki, Lídia Kadlecsik, Attila Gyenesei, Róbert Herczeg, Judit Simon

## Abstract

**Introduction:** SARS-CoV-2 infections have very different clinical manifestations and anti-SARS-CoV-2 immunisation may also trigger very different levels and length of protection. While (re)infection after previous COVID-19 illness or following vaccination are known, their impact and the optimal timing of any booster vaccination is currently debated. International evidence about potential underlying immune response differences remains limited and is currently not available in Hungary.

**Methods:** We prospectively investigated the magnitude of immune responses to infection or immunisation, their over-time changes and the occurrence of new infections through anti-SARS-CoV-2 IgG levels and the association with selected individual and clinical parameters in two voluntary cohorts of healthcare workers at a public teaching hospital in a real-world longitudinal cohort study in Hungary. In the first cohort, the anti-nucleocapsid IgG levels of 42 health care workers (female: 100%) with SARS-CoV-2 infection were followed-up over 8 months between June 2020 and February 2021. Beyond the change in immune response, associations with age, selected existing chronic conditions, blood type and severity of symptoms were investigated. In the immunised cohort, anti-spike-RBD protein IgG levels of 49 health care workers (female: 73%) with no prior COVID-19 infection were monitored up to 4 months following initial immunisation with BNT162b2 vaccine between December 2020 and April 2021. Statistical analyses included median analysis, linear regression, ANCOVA, Kruskal-Wallis test and Skillings-Mack test for block designs as relevant.

**Results:** Within the infected cohort, the median time of anti-SARS-CoV-2 IgG level reduction below the positive test cut-off was 6 months. First month IgG levels were on average the highest among those in illness severity category 4, but the difference to less severe categories was not statistically significant. Higher age was associated with higher IgG levels. Within the immunised cohort, the anti-SARS-CoV-2 spike-RBD protein IgG levels increased 25-fold between the first and second immunisations, significantly decreased to 33% of the peak level after 90 days, and had an overall negative tendency with older age and male sex. IgG monitoring revealed 17% (7/42) and 14% (7/49) new infections in the infected and the immunised cohorts, respectively, all symptomless.

**Discussion:** Our study is the first to investigate the level, change and associations of anti-SARS-CoV-2 IgG immune response in infected or immunised healthcare workers in Hungary. It provides further evidence about the significantly declining IgG protection through initial infection beyond 6 months. While immunisation with mRNA vaccination shows a similar pattern of reduction in protection, IgG levels remained within the positive range at 4 months. The observed rate of 15% new, asymptomatic infections and their potential broader impacts call for further investigations. Overall, our findings are confirmative of the effectiveness of vaccination to prevent illness, recent considerations for booster vaccination beyond 6 months, and indicate the potential benefit of anti-SARS-CoV-2 IgG monitoring for optimisation.

## Introduction

The novel SARS-CoV-2 is a single-stranded positive-sense RNA coronavirus. It has four structural proteins, the nucleocapsid protein (N), the membrane protein (M), the spike protein (S), and the envelope protein (E). From these, the receptor-binding domain (RBD) of subunit S1 attaches to the host cell’s angiotensin-converting enzyme 2 receptor and enters the cell’s cytoplasm, playing a central role in the infection. [1] Human-to-human transmission may happen via droplets, infected body fluids and direct or indirect contact both in the case of asymptomatic and symptomatic infections. Diagnosis of the viral infection is done with real-time polymerase chain reaction (PCR) tests from nasopharyngeal samples [2].

The first SARS-CoV-2 infections were identified in Wuhan, China in December 2019, while in Hungary the first case was diagnosed on 4 March 2020. One week later, on the 11 March 2020, the World Health Organisation (WHO) declared the outbreak a pandemic. The infection with SARS-CoV-2 is responsible for the COVID-19 respiratory illness and may cause very various symptoms and run with different illness severities. The reported case fatality rate of COVID-19 in Europe lies between 0.2% in Germany and 7.7% in Italy and varies over time, according to geographical location, level of testing and reporting with an increased risk of dying with older ages and with underlying health conditions such as respiratory illness, diabetes mellitus, and cardiovascular disease [3].

While currently no specific medical therapy of COVID-19 exists, anti-SARS-CoV-2 vaccines for the prevention of severe COVID-19 illness with the spike protein as main target have been available since late 2020. There are also diverse highly sensitive and specific automated serological tests available for the measurement and monitoring of humoral immune response induced either by infection in the form of anti-nucleocapsid IgG or by vaccination in the form of anti-spike-protein-RBD IgG. Routine tests for the measurement of cellular immunity against SARS-CoV-2 are being developed. [4, 5, 6]

Among the currently approved vaccines, all but one require an initial dose followed by a second dose given 3-12 weeks later. A study conducted in the USA regarding the effectiveness of mRNA vaccines for the prevention of severe illness among health care workers under real-world circumstances showed an overall 90% effectiveness 14 days after the second vaccine [7]. The rate of effectiveness after two vaccinations with BNT162b2 was 95% [8], and was confirmed at 85% against the more infectious and severe B.1.1.7 virus mutation [9]. Although the rate of protection is lower with only one vaccine, the level of immune response induced by a single dose of vaccine was found the same for those with previous COVID-19 infection as for others by two doses [10, 11].

Longevity of the induced immune response, the rate of new infections and the risk of further spreading following previous illness or immunisation are currently major research questions with limited relevant evidence [12]. According to the ongoing UK SIREN study, the rate of protection against new infections among healthcare workers was 83% five months after a previously diagnosed COVID-19 illness and found a less than 1% reinfection rate, mostly asymptomatic [13]. Other studies reported that IgG levels became negative after 6-8 months, however, the level of cellular immunity - especially B-cell activity – seems to remain or even increase with time [14, 15, 16]. A recent meta-analysis found the relative risk of spreading the disease further 42% less for asymptomatic cases in comparison to symptomatic cases [17].

Hungary is the first EU country starting vaccination of healthcare workers with two doses of BNT162b2 21 days apart at the end of 2020, and one of the frontrunners in Europe in terms of vaccination rate. Evidence related to the level and longevity of induced immune response following SARS-CoV-2 infection or immunisation, and their associations with specific individual or clinical factors is, however, so far not available.

Early on in the pandemic, our molecular diagnostic laboratory, a national centre for viral hepatitis diagnostics located within a public teaching hospital in Hungary initiated a prospective real-world longitudinal cohort study to investigate the magnitude of humoral immune response to SARS-CoV-2 infection, eventually to anti-SARS-CoV-2 immunisation, their changes over time, the occurrence of new infections, and their associations with selected individual and clinical parameters through the regular monitoring of anti-SARS-CoV-2 IgG levels in two cohorts of healthcare workers.

## Materials and methods

### Participants and data collection

In cohort 1, volunteering health care workers with previously diagnosed SARS-CoV-2 infection were invited to monthly measurement of their anti-nucleocapsid IgG levels following recovery and monitored over 8 months between June 2020 and February 2021. In cohort 2, anti-spike-RBD protein IgG levels of volunteering health care workers with no prior COVID-19 infection were monitored up to 4 months following initial immunisation with BNT162b2 (Pfizer/BioNTech) vaccine between December 2020 and April 2021.

Recruitment for both cohorts was through the standard internal mailing list of the local hospital over 2 weeks and 1 week for cohorts 1 and 2, respectively. Participants had to be over 18 years, employed by the hospital, currently not on high-dose immune suppressive therapy, and willing and able to give written consent to participate in the study.

For cohort 1, inclusion and first blood sample collection were based on negative PCR test following previous confirmed diagnosis of SARS-CoV-2 infection by positive PCR test. Individual and clinical parameters were collected via a self-completion questionnaire, including information on age, blood group, pre-existing health conditions such as diabetes or autoimmune disorders and related medication use, and type of department. Severity of COVID-19 symptoms were grouped into six categories: 0=no symptoms, 1=mild symptoms, 2=moderate symptoms, 3=severe symptoms (no hospitalisation), 4=hospitalisation with or without intensive care, 5=intensive care with ventilation.

For cohort 2, inclusion and first blood sample collection was prior to receiving the second vaccine dose 21 days after the first vaccine dose upon confirmation of no previous infection through a negative anti-nucleocapsid IgM/IgG test. Information on age and type of department were available from routine records. Blood sample taking was repeated in relation to the first vaccination on days 21+7, 21+7+60 and 21+7+90.

The study was approved by the Local Institutional Committee of Science and Research Ethics (Nr. 16/20200723).

### Laboratory methods

For cohort 1, anti-SARS-CoV-2 nucleocapsid IgG levels were measured with chemiluminescent microparticle immunoassay method (CMIA) using the semi-quantitative SARS-CoV-2 IgG/IgM (Abbott) and Architect i2000SR immunoassay analyser (Abbott). Positive diagnostic levels were defined as >1.4 s/c. [4, 6, 18]

For cohort 2, anti-SARS-CoV-2 spike-RBD levels were measured with chemiluminescent microparticle immunoassay method (CMIA) using the quantitative SARS-CoV-2 IgG II Quant Assay (Abbott) and Architect i2000SR immunoassay analyser (Abbott). Positive diagnostic levels were defined as >50 AU/ml. [5, 18]

## Statistical analysis

For cohort 1, change in immune response (positive to negative) over time, associations between the initial levels of anti-SARS-CoV-2 nucleocapsid IgG (month 1) with age, existing selected chronic conditions, blood type and severity of symptoms were investigated. For cohort 2, the change in the level of anti-SARS-CoV-2 spike-RBD protein IgG over time and the associations between the initial levels of immune response following first vaccination (day 21) and second vaccination (day 21+7) with age and gender were analysed.

Statistical analyses were censored for vaccinations in cohort 1 and for new infections in both cohorts. For cohort 1, missed monthly IgG level measurement data were interpreted in light of the next available measure assuming a linear change between the measurements available before and after the missing value.

For cohort 1, median analysis for change in immune response over time, linear regression to examine the association between the level of IgG and age, and Kruskal-Wallis test for association with symptom severity were used. For cohort 2, ANCOVA was applied where age and sex were taken into account to model the relationship with the level of IgG, and Skillings-Mack test with block designs was used for the longitudinal analysis of anti-spike-RBD protein IgG levels. Statistical significance was defined as p≤0.05.

Statistical analyses were done using R statistical software v3.6.1 (R Core Team, 2019) applying and lm function and the Skillings.Mack package (v1.10) [19, 20, 21].

## Results

Cohort 1 comprised of 42 female participants (100%), mean age 47 years (s.d.: 8). Blood type distributions were A/B/AB/0=17/10/6/9 (41%/24%/14%/21%) and Rh+/-=37/5 (88%/12%). Underlying health conditions known to influence immune reactions such as autoimmune diseases and diabetes mellitus were identified in 6 and 4 (Type II/I: 3/1) participants, respectively. All those with an autoimmune disease received only low dose immune suppressive medications deemed eligible. Symptom severity distribution according to categories 0/1/2/3/4/5 was n=4/5/10/14/9/0 (10%/12%/24%/33%/21%/0%). Mean anti-SARS-CoV-2 nucleocapsid IgG level in month 1 was 6.68 s/c (s.d.: 2.13) and levels were positively associated with age (+0.085; p=0.035). Category 4 (the highest category in our sample) had the highest mean level of anti-SARS-CoV-2 nucleocapsid IgG with 7.85 s/c (s.d.: 0.59) at month 1, although the difference to less severe categories was not statistically significant. (**Fig 1**)

**Figure 1.**
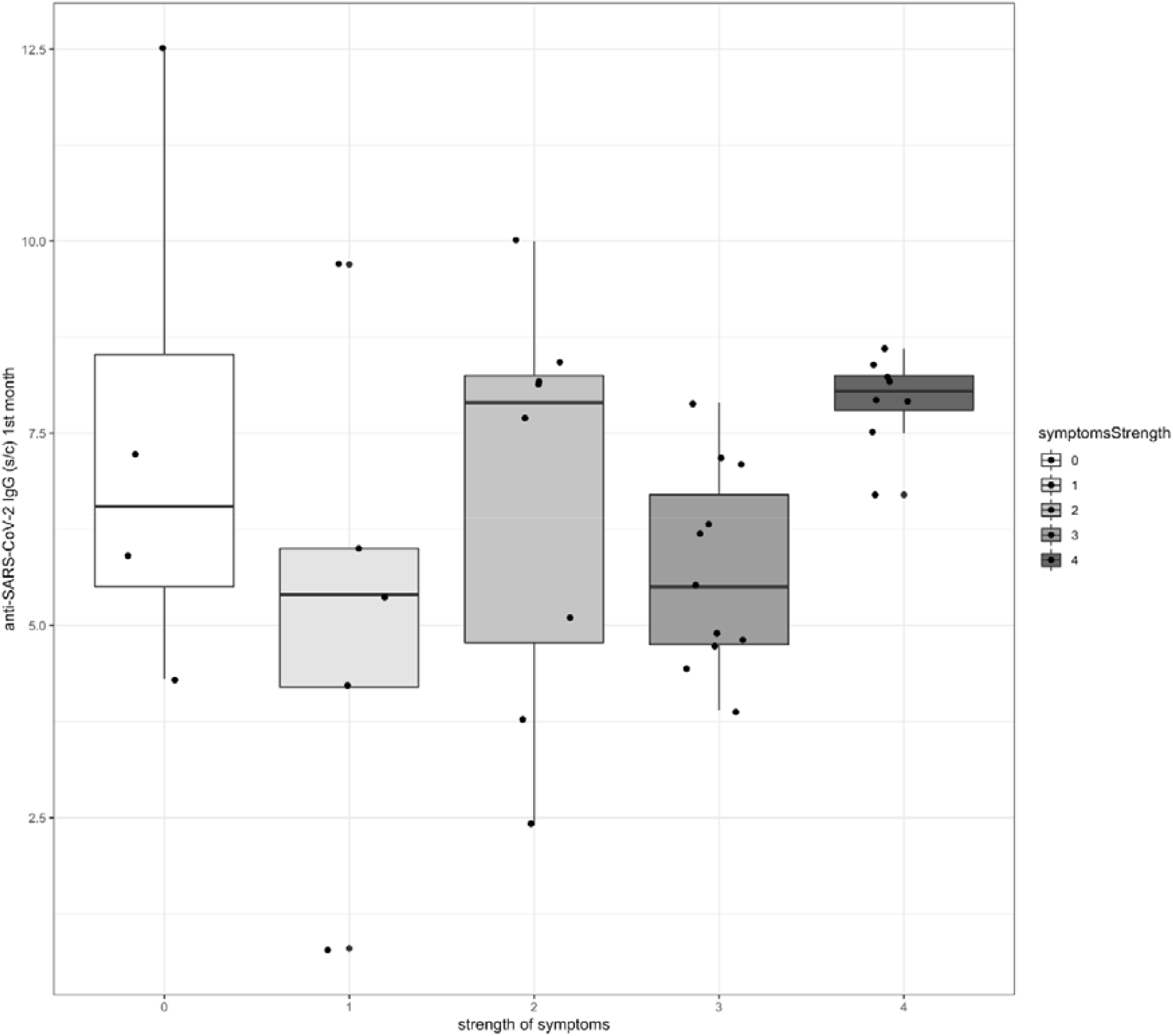
Association between anti-SARS-CoV-2 nucleocapsid IgG levels in month 1 and symptom severity in Hungarian healthcare workers

Seven out of the 42 participants in cohort 1 had a sudden increase in their anti-SARS-CoV-2 nucleocapsid IgG during monitoring (M3/M4/M5: n=4/1/2) indicating reinfections and were removed from further analysis. All were asymptomatic, six of them worked at high risk departments (COVID ward, infectious diseases). Other ten participants were censored due to vaccination (M6/M7/M8: n=3/6/1). Regarding the remaining participants, random missingness in single measurement data with informative value was 11%. Minimum time for drop in the anti-SARS-CoV-2 nucleocapsid IgG level below the non-positive cut-off was 3 months (n=1), four persons retained their positive levels at month 8. Median time of change to non-positive values was 6 months.

Cohort 2 had 49 participants, 76% females (male/female: 13/37), mean age was 53 years (s.d.: 12). Mean anti-SARS-CoV-2 spike-RBD protein IgG level was 717 AU/ml (s.d.: 592) following first vaccination (day 21) and 17987 AU/ml (s.d.: 11166) following second vaccination (day 21+7), a 25-fold increase (**Table 1**). At both time points the levels of IgG immune response were negatively influenced by age (ANCOVA: -12, p=0.201; -317, p=0.074) and male sex (ANCOVA: -1045, p=0.162; -130086, p=0.347), but these associations did not reach statistical significance.

**Table 1.** Anti-SARS-CoV-2 nucleocapsid IgG levels following immunisation of healthcare workers in Hungary

|  | Age<br>mean<br>(s.d.) | 21 days after 1<sup>st</sup><br>vaccination<br>(AU/ml) | 7 days after<br>2<sup>nd</sup> vaccination<br>(AU/ml) | 7+60 days after<br>2<sup>nd</sup> vaccination<br>(AU/ml) | 7+90 days after<br>2<sup>nd</sup> vaccination<br>(AU/ml) |
| --- | --- | --- | --- | --- | --- |
| female | 51.01<br>(9.59) | 831<br>(602) | 20055<br>(11470) | 9847<br>(6578) | 6907<br>(8453) |
| male | 58.07<br>(16.45) | 400<br>(440) | 12259<br>(8162) | 4971<br>(2905) | 3190<br>(2022) |
| all | 52.87<br>(12.03) | 717<br>(592) | 17987<br>(11166) | 7313<br>(3953) | 4434<br>(2364) |

All values are represented as mean and standard deviations (s.d.).

Seven out of the 49 participants experienced a sudden increase in their anti-SARS-CoV-2 spike-RBD protein IgG level during further monitoring, six of them at measurement point 3 (day 21+7+60) and one at measurement point 4 (day 21+7+90) indicating COVID-19 infections and were removed from further analysis. All seven persons worked at high risk departments (COVID ward, otorhinolaryngology, PCR laboratory, endoscopy), but had no symptoms.

The level of IgG immune response dropped by 59% between measurement points 2 (days 21+7) and 3 (day 21+7+60), and by 20% between measurement points 3 (day 21+7+60) and 4 (day 21+7+90). Results showed statistically significant differences between all 4 measurement points (Skillings-Mack = 127.412, p<0.0001). (**Fig 2**)

**Figure 2.**
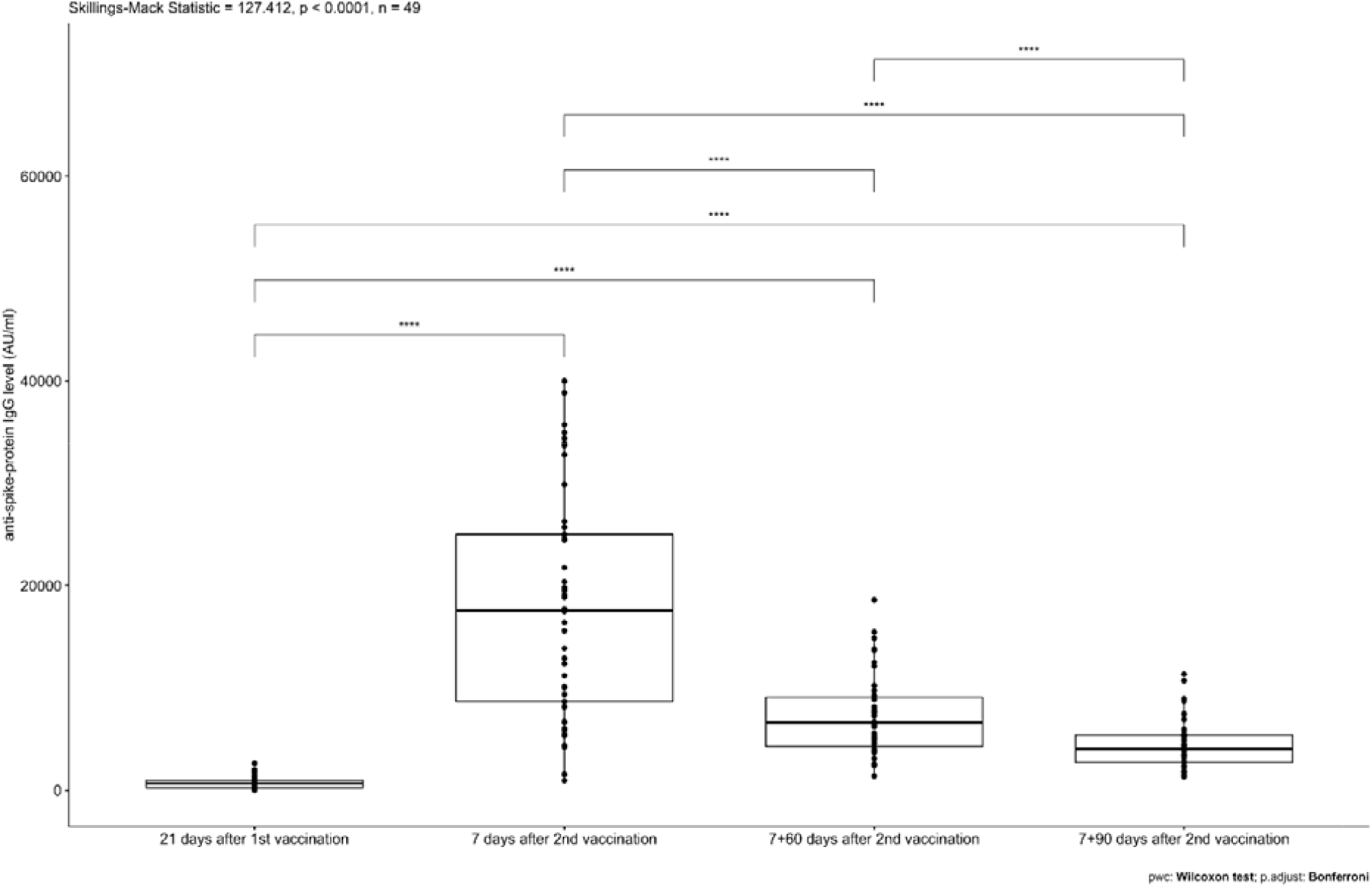
Change in the level anti-SARS-CoV-2 spike-RBD protein IgG over time in Hungarian healthcare workers

## Discussion

We prospectively investigated the magnitude of immune response to infection or immunisation, their over-time change and the occurrence of new infections through anti-SARS-CoV-2 IgG levels and their association with selected individual and clinical parameters in two voluntary cohorts of healthcare workers at a public teaching hospital in a real-world longitudinal cohort study in Hungary.

Within the infected cohort, the median time of anti-SARS-CoV-2 IgG level reduction below the positive test cut-off was 6 months. Although first month IgG levels were on average the highest among those in illness severity category 4, the difference across categories was not statistically significant. Higher age was associated with higher IgG levels. Our findings partially confirm the results of Havervall and colleagues who followed-up 2300 previously SARS-CoV-2 infected healthcare workers in Sweden and found the average time for the reduction of anti-SARS-CoV IgG below the positive cut-off point between 4-6 months [22]. They also found the association between higher immune response levels and more severe symptoms statistically significant similar to the study by Long et al. conducted earlier on 37 asymptomatic persons from China [4, 22]. The lack of statistical significance between symptom severity and level of immune response in our study is potentially linked to our limited sample size.

Within the immunised cohort, the anti-SARS-CoV-2 spike-RBD protein IgG levels increased 25 times between the first (day 21) and second immunisation (day 21+7), and significantly decreased to 33% of the peak level at 4 months (day 21+7+90) confirming the importance of the second immunisation in the development of the humoral immune response. The level of anti-SARS-CoV-2 spike-RBD protein IgG had a negative tendency with older age and male gender. Although the association in our study did not reach statistically significance, this is most likely due to the limited sample size. A similar result was reported by Elliot et al. in the UK REACT study which found significant negative association between vaccine-induced immune response measured by lateral flow immunoassay test to detect IgG against SARS-CoV-2 spike protein and age. Among those who received Pfizer/Biontech vaccine, merely 35% of those above the age of 80 years had a positive IgG level after the first vaccination and 88% after the second vaccination as opposed to 95% and 100% of those under the age of 30 years. [23]

Altogether we identified 17% (7/42) and 14% (7/49) symptomless new infections in the infected and the immunised cohorts, respectively. Our study is confirmative about the effectiveness of the vaccination to prevent illness and of other existing international evidence about the possibility of (re)infection despite positive anti-SARS-CoV-2 immune response, although, it shows a potentially higher rate of (re)infections in comparison to the UK SIREN study [13]. This higher rate of (re)infections based on the sudden increase in IgG levels should be interpreted within the given study context where testing of healthcare workers in the hospital was conducted dominantly targeted rather than as part of systematic screening. Since all cases were asymptomatic and IgG measurements happened in monthly intervals, it was not feasible to get additional PCR test confirmation.

Our study is the first to investigate the level and over-time change of anti-SARS-CoV-2 IgG immune response and its associations with individual and clinical parameters in infected (cohort 1) or immunised (cohort 2) healthcare workers in Hungary. It has been conducted applying robust methods under real-world circumstances with volunteering participants and has the longest possible monitoring period concerning independent investigations of vaccine-induced IgG immunity within the EU. It provides further evidence about the significantly declining IgG protection through initial infection beyond 6 months. While immunisation with mRNA-vaccination showed a similar pattern of reduction in protection, IgG levels remained within the positive range at the current maximum follow-up period of 4 months.

Our findings are in line with the limited current international evidence in the field, but due to our comparatively small sample size when considering studies from the UK, Sweden or China, the statistical power of our analyses remains low. Due to the voluntary nature of our recruitment, we have also experienced a major imbalance in the sex distribution of our cohort 1 where all included participants are female. This limits the validity of our findings about IgG immune response following SARS-CoV-2 infection to women only and does not allow the investigation of any associations by sex. On the other hand, cohort 1 represents 2.5% of the workers actively involved in clinical care at our hospital (n=1680) and 1.4% of the total number of employees (n=2956). Finally, our study focused on the monitoring of anti-SARS-CoV-2 IgG levels and have no additional data about the cellular immunity of our participants.

## Conclusions

Overall, our findings are confirmative of the emerging plans about a third booster vaccination to prevent COVID-19 beyond 6 months. The observed rate of approximately 15% new, asymptomatic infections mostly among healthcare workers with positive anti-SARS-CoV-2 IgG levels working in high risk areas confirms the effectiveness of the vaccination to prevent illness. On the other hand, this information calls for further considerations about the necessity of sustained preventive measures and suggests great potential benefits of targeted IgG level monitoring of healthcare workers in high risk areas until at least we have better understanding of the infectious potential of these cases.

Monitoring of cohort 2 is ongoing in order to be able to establish the median time of change of anti-SARS-CoV-2 IgG immune response below the positive cut-off. Future findings should contribute to the optimised timing of planned booster vaccinations and may highlight further potential benefits of regular anti-SARS-CoV-2 IgG monitoring of healthcare workers.

## Data Availability

Study data are available upon request from the corresponding author.

## Acknowledgments

The authors would like to thank the expert support of Prof. emer. Kovács L. Gábor, Member of the Hungarian Academy of Sciences, University of Pécs, Pécs, Hungary.

## Author contributions

Conceptualization: JG; data collection and laboratory analysis: KSz, EB, LK supervised by JG; statistical analysis: RH, AGy; interpretation of results: JG, JS; writing—original draft preparation: JG, JS; revision and approval of manuscript: all authors.

